# COVID-19 Susceptibility, Mortality, and Length of hospitalization based on age-sex composition: Evidence from Davao Region Philippines

**DOI:** 10.1101/2021.06.20.21259222

**Authors:** Roel F. Ceballos

## Abstract

The coronavirus disease is spreading continuously worldwide with an unprecedented amount of impact on every human society. In order to reduce the risks of infections and mortality, several interventions such as mobility restrictions for different age groups and vaccination prioritization programs are implemented in the Philippines. Identifying age-sex composition with greater susceptibility, longer hospitalization, and higher fatality is useful to guide the targeted intervention and establish risk stratification for patients infected with COVID-19 within communities and localities. Furthermore, it is also helpful in the allocation of medical resources and assessment of vaccination priority. We analyzed the COVID-19 data provided by the Davao Center for Health Development of the Department of Health Davao Region in the Philippines. The dataset contains records of COVID-19 cases reported from March 2020 to April 2021. Methods that were used include descriptive statistics, graphical presentations, and nonparametric statistical methods. The study reveals that male children and female senior citizens are the most susceptible age-sex composition while male senior citizen is the subgroup with the highest case fatality and mortality. Furthermore, regardless of sex groups, the senior citizen is the subgroup with the longest hospitalization. Susceptibility due to exposure should be included as a criterion in determining the age-sex compositions for vaccination priority against COVID-19 and other potentially deadly viruses. Further, Proper planning and allocation of medical resources for the elderly should be prioritized in the provincial levels.

## INTRODUCTION

The coronavirus disease 19 (COVID-19) is a highly transmittable disease caused by severe acute respiratory syndrome coronavirus 2 (SARS-CoV-2), which emerged in Wuhan, China, and spread around the world^1^. On January 30, the World Health Organization announced a Public Health Emergency of International Concern (PHEIC) for the COVID-19 outbreak^2^. As of June 3, 2021, the virus has spread to almost 200 countries and territories, including 26 cruise and naval ships^3^. In the Philippines, several interventions such as restrictions of mobility and vaccination prioritization programs according to age groups are implemented to reduce the risk of infection and mortality. Identifying age-sex composition with higher susceptibility and fatality will enable health experts and policymakers to take more effective surveillance and targeted interventions to minimize the pandemic’s adverse effects. Based on the hospital data, earlier epidemiological studies showed that age and sex are two of the most important factors that influence the susceptibility and severity of COVID-19^4,5,6^. However, these studies have mainly concentrated on large economies, and little is known whether the reported age-gender-dependent patterns exist in low-income/middle countries, explicitly considering data within community levels and local government units.

This study aims to determine the composition of age and gender that are highly susceptible to COVID-19 infection, with longer hospitalization and at higher mortality risks, to guide the targeted intervention and establish risk stratification for patients infected with SARS-CoV-2 in Davao Region, the most populous in the whole Mindanao Island and contains within its territory the City of Davao which is the largest city in the Philippines based on land area.

## METHODS

### Study Design and Data

A retrospective design was employed in this study involving COVID-19 positive cases in Davao Region from March 2020 to April 2021. The data was provided upon request by the Davao Center for Health Development of the Department of Health Davao Region located at J.P. Laurel Avenue Davao City, Philippines. It comprised a total of 24,295 cases containing the following information, age, sex, province, date of admission to either hospital or quarantine facility, the status of whether recovered or died or still admitted at the time of data retrieval. To calculate mortality rates, the researcher retrieved the population by age-sex composition from the publication of the Philippine Statistics Authority, a government agency responsible for the development and conduct of population census in the country. The retrieval and use of both the COVID-19 data and population data of the Davao Region comply with the 2012 Data Privacy Act in the Philippines because these data sets do not contain a personal identifier that may expose the identity of individuals.

### Statistical Analysis

Statistical analysis was carried out using the R Programming language. It is a free software environment for statistical computing and graphics supported by the R Foundation for statistical computing^7^.In order to determine and compare the susceptibility and mortality of the different age and sex combinations, the following were used: frequency, percentage, bar charts, nonparametric statistical procedures such as Chi-square test and correlation plot of Chi-square residuals. The male to female sex ratio was also used to compare the susceptibility and mortality of the different age groups by the province in the Davao Region. Case fatality ratio was also computed in addition to the methods used to measure mortality of COVID-19 in the Region. Lastly, median, interquartile range, and Kruskal-Wallis test was used to compare the hospitalization of COVID-19 patients in the Davao Region.

## RESULTS

### Susceptibility by Sex and Age composition

One of the aims of this research is to determine the age-sex combinations that are highly susceptible to COVID-19 infection upon exposure. Table 1 shows the number and percentage of cases for the different age-sex compositions from March 2020 to April 2021 in Davao Region, Philippines. The age group is categorized into four: children (0 to 12 years old), teens (13 to 19 years old), Working adults (20 to 59 years old), and Senior Citizens (60 years old and above). The highest reported cases are in the Working Adults group for both men and women, which is 74% and 73.8%, respectively. The result is followed by senior citizens (12% and 13%).

**Table 1.** COVID-19 cases by sex and age from March 2020-April 2021 in Davao Region

| Sex Group | Age group |
| --- | --- |

|  |  | *Children* | *Teens* | *Working adults* | *Senior Citizens* |
| --- | --- | --- | --- | --- | --- |
| *Men* | Cases, n | 870 | 838 | 9029 | 1460 |
|  | (%) | (7.1) | (6.9) | (74.0) | (12.0) |
| *Women* | Cases n(%) | 732 | 854 | 8932 | 1589 |
|  | (%) | (6.1) | (7.1) | (73.8) | (13.1) |

The proportions of COVID-19 cases in women are higher than 50% among teens and senior citizens, while male cases are higher than 50% among children and working adults. The difference in proportions of COVID-19 cases among age-sex combinations is significant (X2 test, p-value<0.01, Fig. 1a), which implies that some subgroups are more susceptible than others. To further investigate these differences in susceptibility, the residuals of the X2 test are presented using a correlation plot. The correlation plot reveals that among COVID-19 cases in Davao Region, male children and female senior citizens are more susceptible to COVID-19 infection than other age and sex combinations (Fig.1b).

**Figure 1.**
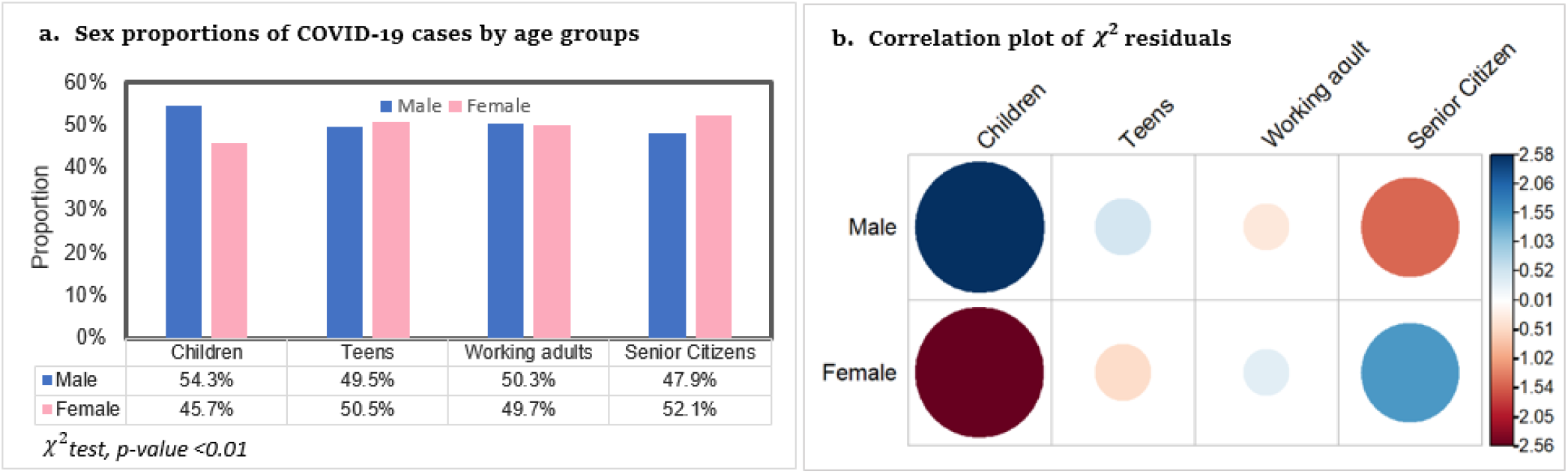
COVID-19 cases in Davao Region by sex and age groups.

We are interested to know if the same pattern exists at provincial levels. For this purpose, we present in Table 2 the male-to-female sex ratio of COVID-19 cases for the different age groups by province in the Davao Region. Davao Region is composed of five provinces, namely: Davao del Sur, Davao del Norte, Davao de Oro, Davao Oriental and Davao Occidental. It is observed that across all provinces in Davao Region, male children have greater susceptibility than female children (SR: 1.03 to 1.58). A result that is consistent with the correlation plot in Figure 1b. Furthermore, higher susceptibility of female senior citizens is observed in Davao de Oro and Davao Oriental, while a close male-to-female ratio is observed in Davao del Sur and Davao Del Norte.

**Table 2.** Sex Ratio of COVID-19 cases per 100,000 population in Davao Region

| Province | Age group |  |  |  |
| --- | --- | --- | --- | --- |
|  | *Children* | *Teens* | *Working adults* | *Senior Citizens* |
| Davao del Sur | 1.12 | 1.09 | 1.00 | 1.07 |
| Davao del Norte | 1.03 | 0.78 | 0.80 | 1.04 |
| Davao de Oro | 1.58 | 0.79 | 0.81 | 0.92 |
| Davao Oriental | 1.03 | 0.77 | 0.77 | 0.80 |
| Davao Occidental | 1.21 | 1.11 | 1.29 | 1.22 |

### Length of hospitalization

Another aim of this study is to examine the length of hospitalization among subgroups based on age-sex compositions. First, we present the length of hospitalization for the recovered group in Figure 2, and second, we present the length of hospitalization for the mortality group or those patients who died in Figure 3. Our analysis revealed that similar patterns on the differences of hospital days among age groups are observed in both sex groups wherein senior citizens have the longest hospitalization followed by working adults. In addition, the length of hospitalization for females is statistically different for all age groups (p-value<0.01) (Fig.2b). The result is almost the same for the males, except that the length of hospitalization for children and teens are not statistically different (Fig.2a)

**Figure 2.**
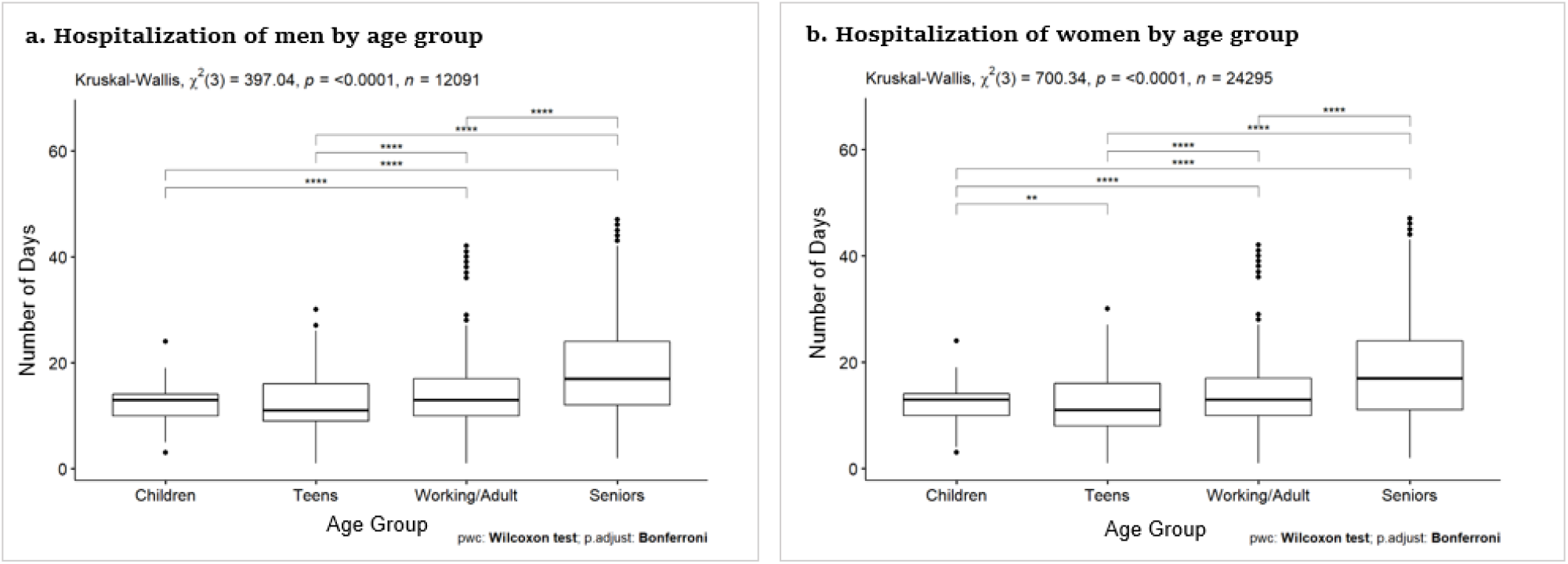
Hospitalization for recovered COVID-19 cases.

**Figure 3.**
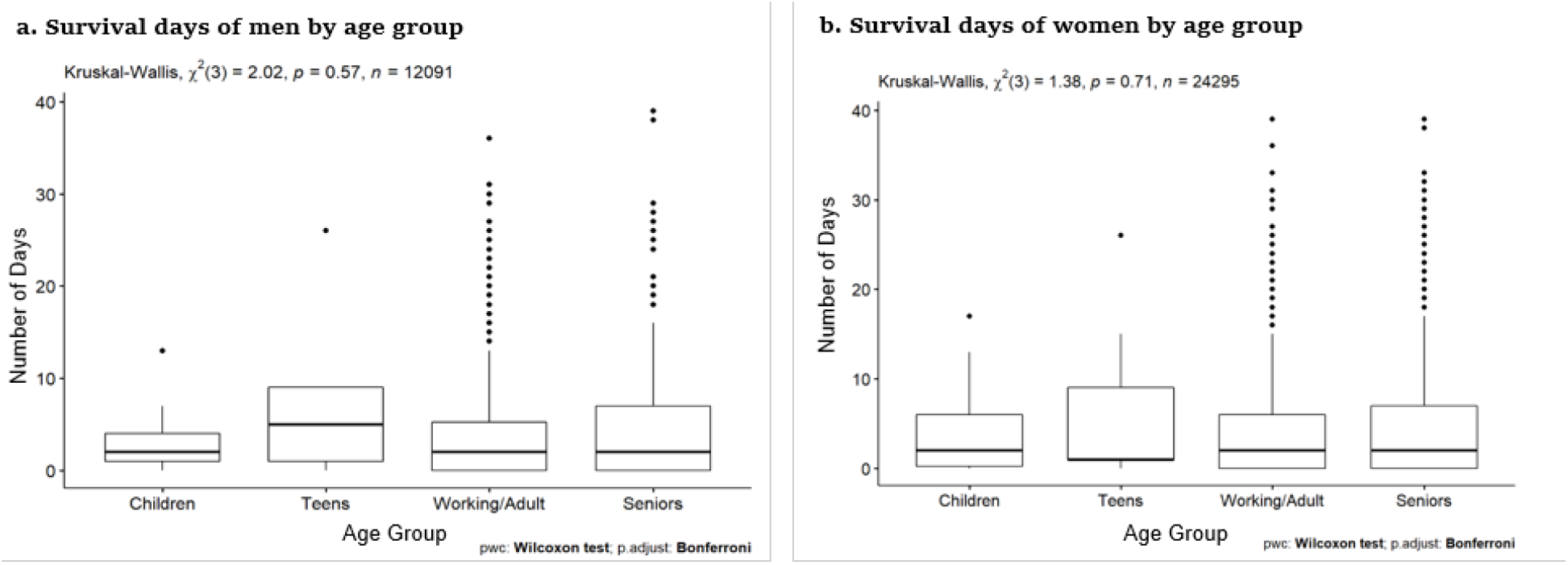
Hospitalization for mortality group.

Furthermore, the length of hospitalization for the mortality group were examined using the Kruskal-Wallis test (Fig.3a for men and Fig. 3b for women). Results revealed no significant difference in the hospital days for the different age groups in both men and women.

### Case Fatality Rate and Mortality

We are also interested in examining the case fatality and mortality of the different combinations of age and sex. Table 3 shows that most reported deaths are among senior citizens, 52% for men and 61% for women. The result is followed by working adults, 45% for men and 36% for women. Similar patterns of case fatality and mortality rates are observed for both groups in which senior citizens have the highest rate.

**Table 3.** Case fatality rates and Mortality rates of COVID-19 cases in Davao Region

| Sex Group | Age group |  |  |  |
| --- | --- | --- | --- | --- |
|  | Children | Teens | Working adults | Senior Citizens |
| *Men* |  |  |  |  |
| Deaths, n | 17 | 5 | 268 | 312 |
| (%) | (2.8) | (0.8) | (44.5) | (51.8) |
| cCFR % | 2.0 | 0.6 | 3.0 | 21.4 |
| Mortality/100,000 population | 1.23 | 0.65 | 9.91 | 71.15 |
| *Women* |  |  |  |  |
| Deaths n(%) | 9 | 4 | 144 | 245 |
| (%) | (2.2) | (1.0) | (35.8) | (60.9) |
| cCFR % | 1.23 | 0.47 | 1.61 | 15.42 |
| Mortality/100,000 population | 0.65 | 0.52 | 5.33 | 55.87 |

Furthermore, males’ crude case-fatality ratio is significantly higher than females in working adults and senior citizens. Mortality per 100,000 population directly caused by COVID-19 among males is significantly higher than among females for all age groups except for teens (proportion test, p-value<0.01 Fig.4).

**Figure 4.**
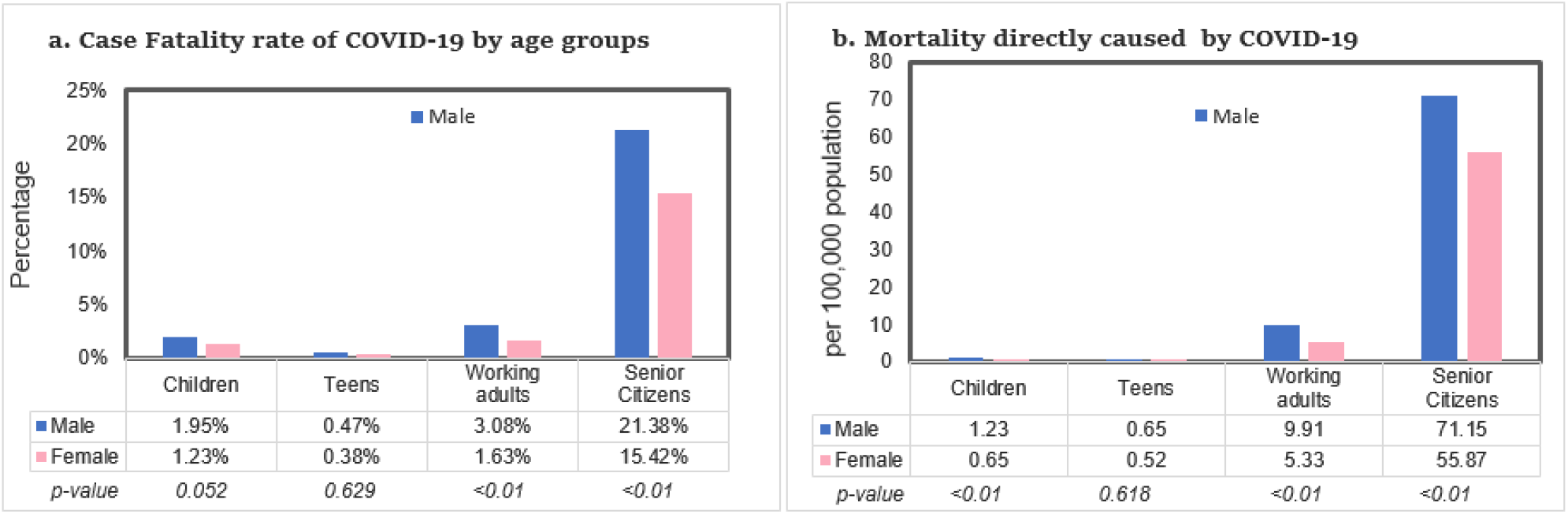
COVID-19 cCFR and mortality in Davao Region by sex and age groups.

Furthermore, we investigated these findings by considering the male-to-female Sex Mortality Ratio in each age group by province. We are interested to know if the same pattern exists at provincial levels. Table 4 shows that the sex mortality ratio of COVID-19 cases for each province in the Davao Region is higher for men, ranging from 1.01 to 1.54 in different provinces, which implies that indeed male senior citizens have higher mortality.

**Table 4.**
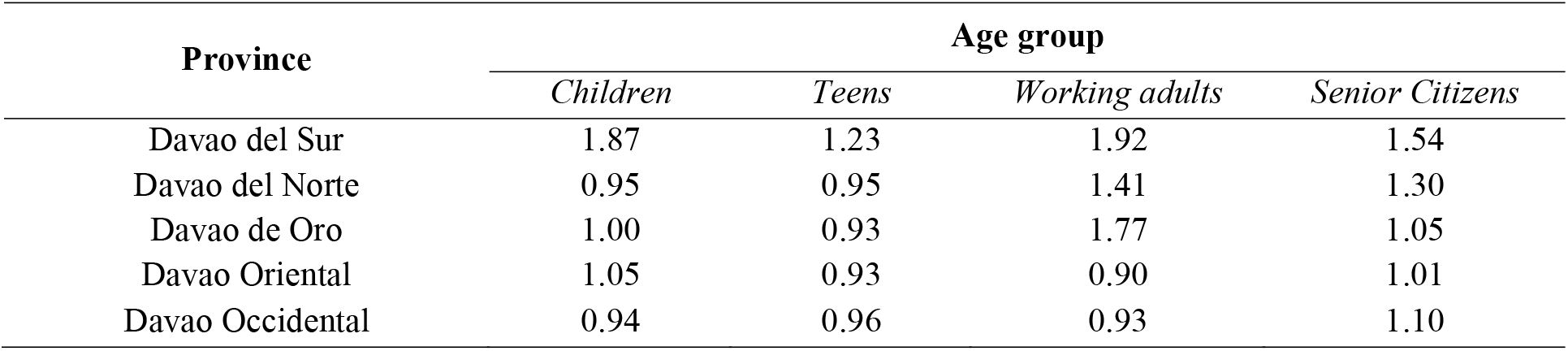
Sex Ratio of Mortality Rates

## DISCUSSION

The purpose of lockdowns and vaccinations is to prevent mortality, preserve the health system’s capacity, and protect the populations most at risk. Therefore, it is imperative to achieve these goals such that the most susceptible population subgroups with longer hospitalization tendency and at higher risk of mortality shall be prioritized by vaccinations and should be considered in policies concerning overall COVID-19 management such as allocation of hospital resources and limitations of mobility. This study aims to identify age and sex composition with greater susceptibility, longer hospitalization, and higher fatality to provide a guide for the targeted intervention and the establishment of risk stratification for patients infected with COVID-19 and allocation of medical resources and assessment of vaccination priority.

Results reveal that the working adults are the most susceptible among the age groups, followed by the senior citizens. The working adult group is composed of frontline health workers and essential workers. Thus, this group has greater exposure to COVID-19 infection because the nature of their work requires them to move around constantly^8^. On the other hand, senior citizens have greater susceptibility, despite having lower exposure. The COVID-19 Inter-agency Task Force for the Management of Emerging Infectious Diseases (IATF) in the Phippines prohibits senior citizens from going around except for buying food and medicine because they are prone to comorbidity and low immune system^9^. Senior citizens have the highest case fatality and mortality rate. It is imperative that the senior citizens should be protected through vaccination to reduce their risk of infection and mortality. As of this writing, frontline health workers and senior citizens have already been vaccinated in Davao Region, well at least for those who are willing.

Furthermore, male children are highly susceptible to COVID-19 infections compared with other age-sex compositions. What may explain this is that generally, males are more vulnerable to infections due to sex-based immunological differences^10^.Another explanation may be is that children below five years old have low immunity and are at higher risk of infections^11^. The good news is that, despite being very susceptible to COVID-19 and other viral infections, children are very much adapted to respond and have a lower risk of mortality than other age groups^12^. In addition, although female senior citizens came second in the overall susceptibility of different age-sex compositions, male senior citizens have a greater risk of mortality. Two statistical reports by the World Bank explains this. First, the general life expectancy of Filipinos is shorter for males (67.2 years) than for females (75.5 years). Second, the overall mortality rate of Filipinos is higher for males (233.697 per 1,000 males) than for females (129.01 per 1000 females) ^13^. Several studies have confirmed the association of male sex to COVID-19 deaths ^14, 15^.

Allocation of hospital resources and provision of medical care for patients infected with COVID-19 is an important aspect of COVID-19 management. Our results reveal that hospital stay is longer for aging patients regardless of sex. The same age groups also have significantly higher case fatality rates and mortality rates. These patterns are observable across all provinces in the Region.

## RECOMMENDATIONS

Susceptibility due to exposure should be included as a criterion in determining the age-sex compositions for vaccination priority against COVID-19 and other potentially deadly viruses. In this way, the working adults whose contribution to spurring the economy during the pandemic will be protected primarily. Furthermore, COVID-19 management protocols and policies should be regularly revisited to address and maintain frontline health workers overall wellbeing and safety.

Proper planning and allocation of medical resources for the elderly should be prioritized in the provincial levels since our results reveal that hospital stay is longer for aging patients regardless of sex and that male senior citizens have greater mortality rates.

## Data Availability

All data used in this study are either provided by the appropriate office upon request or are made freely and publicly available on their publication website.

https://drive.google.com/drive/folders/1xlDT96VlzLyvCmMeDZJb2VY3Lh4w7PCw?usp=sharing

